# Effects of transition programs to adulthood for adolescents and young adults with congenital heart disease: a systematic review with meta-analysis

**DOI:** 10.1101/2023.09.11.23295387

**Authors:** Bo Ryeong Lee, Hyun Young Koo, Sangmi Lee

## Abstract

**BACKGROUND:** The increased survival rate among individuals with congenital heart disease (CHD) has sparked interest in their transition to adult healthcare. Although there is a general agreement on the importance of transition interventions, the empirical evidence supporting them is insufficient. Therefore, this study aimed to conduct a systematic review and meta-analysis of transition interventions for adult healthcare in adolescents and young adults.

**METHODS AND RESULTS:** A literature search was conducted for studies comparing the quantitative effects of transition interventions with control groups, published up to March 15, 2023, in major databases (CENTRAL, Embase, PubMed, Web of Science, CINAHL, KISS, and KMbase), major clinical trial registers, academic journal sites related to the topic, and grey literature databases. Ten studies involving a total of 1,297 participants were identified. Transition interventions proved effective in enhancing disease-related knowledge (Hedge’s g=0.89, 95% CI=0.29-1.48) and self-management (Hedge’s g=0.67, 95% CI=0.38-0.95), as well as reducing loss to follow-up (OR=0.41, 95% CI=0.22-0.77). The certainty of evidence for the estimated values of each major outcome was low or very low.

**CONCLUSIONS:** This study supports the implementation of transition interventions by demonstrating that they can improve patients’ disease knowledge and self-management, while also promoting treatment continuity. However, since the available data on transition interventions for adolescents and young adults with CHD remain limited, the widespread adoption of structured transition interventions in the future may alter the conclusions of this study.

**REGISTRATION:** URL: https://www.crd.york.ac.uk/PROSPERO. Unique identifier: CRD42023399026.

**CLINICAL PERSPECTIVE:** *What Is New?:*

- This systematic review of transition programs for individuals of transitional age with congenital heart disease identified 10 relevant studies.
- Transition programs for adolescents and young adults with congenital heart disease were primarily designed to provide individual education, supplement roles related to anatomical and hemodynamic considerations, manage medications and medical appointments and facilitate communication with healthcare providers.
- The transition programs demonstrated efficacy in enhancing disease-related knowledge and self-management, as well as in reducing instances of loss to follow-up. However, they did not significantly improve disease-related quality of life.

*What Are the Clinical Implications?:*

- Given the ethical and practical considerations that arise from country-specific conditions and environments, research on congenital heart disease transition programs should utilize a feasible study design that incorporates a control group.
- Transition programs should address lifestyle factors that can enhance heart function and alleviate clinical symptoms. After promoting long-term commitment to these programs, it is necessary to assess the effects on disease-related quality of life.
- An execution of a transition program that takes into account the developmental characteristics of the target age group, as well as a transition program involving parents, is necessary. Subsequently, an analysis of the effects of these programs is also required.
- As a result of the transition program, it is necessary to measure and analyze not only process indicators but also outcome indicators that directly reflect an individual’s health (e.g. emergency room visits, hospital admissions, and the status of disease or complications).

---

The discontinuation of follow-up among individuals with congenital heart disease (CHD) predominantly occurs during the transition from pediatric to adult care,^1^ and studies have reported that approximately 26.1% of individuals with CHD at the transitional age discontinue their medical follow-up.^2^ Patients who discontinue their follow-up often do not seek medical attention until their heart failure symptoms have significantly worsened, to the point of being life-threatening in some cases.^3,4^ Therefore, it is of vital importance for individuals with CHD to acquire the necessary knowledge and skills for managing their disease in adulthood at the appropriate time, enabling them to independently manage their condition. Additionally, they should transition from pediatric to adult care to ensure they receive lifelong follow-up care.^5,6^

The process through which individuals with CHD attain independent adult disease management, while simultaneously accomplishing medical, psychosocial, educational, and vocational developmental tasks, is referred to as the transition.^6,7^ Those who undergo a suitable transition process gain an understanding of their disease, make informed decisions regarding necessary disease management and a healthy lifestyle (which includes physical activity, diet, career choices, sexual health, and contraception), and are able to utilize the social resources required for adult disease management.^5,6^ Consequently, heart associations across the globe recommend providing transition programs for adolescents and young adults with CHD.^6^

However, the actual impacts and effectiveness of transition programs provided to individuals with CHD are not well-established. Despite growing interest in these programs and related research, their practical implementation for CHD patients is not yet commonplace,^8,9^ and there is a clear need to establish evidence supporting the use of CHD transition programs. Consequently, this study aimed to systematically review the literature on the effectiveness of transition programs for adolescents and young adults with CHD, comparing their outcomes with control groups. Furthermore, through a meta-analysis of various effects, it is hoped that this study will contribute to the evidence base for transition programs for CHD patients.

## METHODS

### 1. Search for Studies

The literature search was conducted in accordance with the COre, Standard, Ideal search (COSI) model.^10^ On February 26, 2023, keyword searches were executed across seven academic databases: CENTRAL, Embase, PubMed, Web of Science, CINAHL, KISS, and Kmbase. From February 26, 2023, to March 15, 2023, additional searches were carried out in clinical trial databases (ClinicalTrials.gov, ICTRP), on relevant academic journal websites, and through grey literature sources (ERIC, ProQuest, RISS, OPENGREY.EU). The search keywords comprised combinations of the following terms: "adolescent/puberty/young adult/pediatrics/child," "heart defects, congenital/congenital heart malformation," and "patient transfer/continuity of patient care/health transition/transition to adult care/self-management/patient transport/patient care/population dynamics/transition to adult care/self-care”. To enhance the sensitivity of the literature search, no restrictions were imposed on the publication year, publication type, or language of the literature.

### 2. Inclusion and Exclusion Criteria

#### 2.1. Population

The studies included adolescents (aged 10-17 years) and young adults (aged 18-29 years) who had simple, moderate complexity, or severe complexity CHD, or those who had experienced cardiomyopathy since childhood. Studies that involved non-structural cardiac conditions or individuals outside the age range of 10-29 years were excluded.

#### 2.2. Interventions

The experimental group (or exposure group) participated in a transition program that incorporated five key components: an introduction to transition, medical knowledge, living with CHD, self-management, and self-advocacy. The goal of this program was to foster independent adult health management, following the guidelines set forth by the American Heart Association (AHA) (Table 1).^5,6^ The intervention (or exposure) was deemed to have been provided if one or more topics within each component were addressed. The control group (or non-exposure group) either received no intervention or usual care. This control group comprised individuals who had not participated in a structured transition program or received any other form of intervention, as defined in the literature.

**Table 1.**
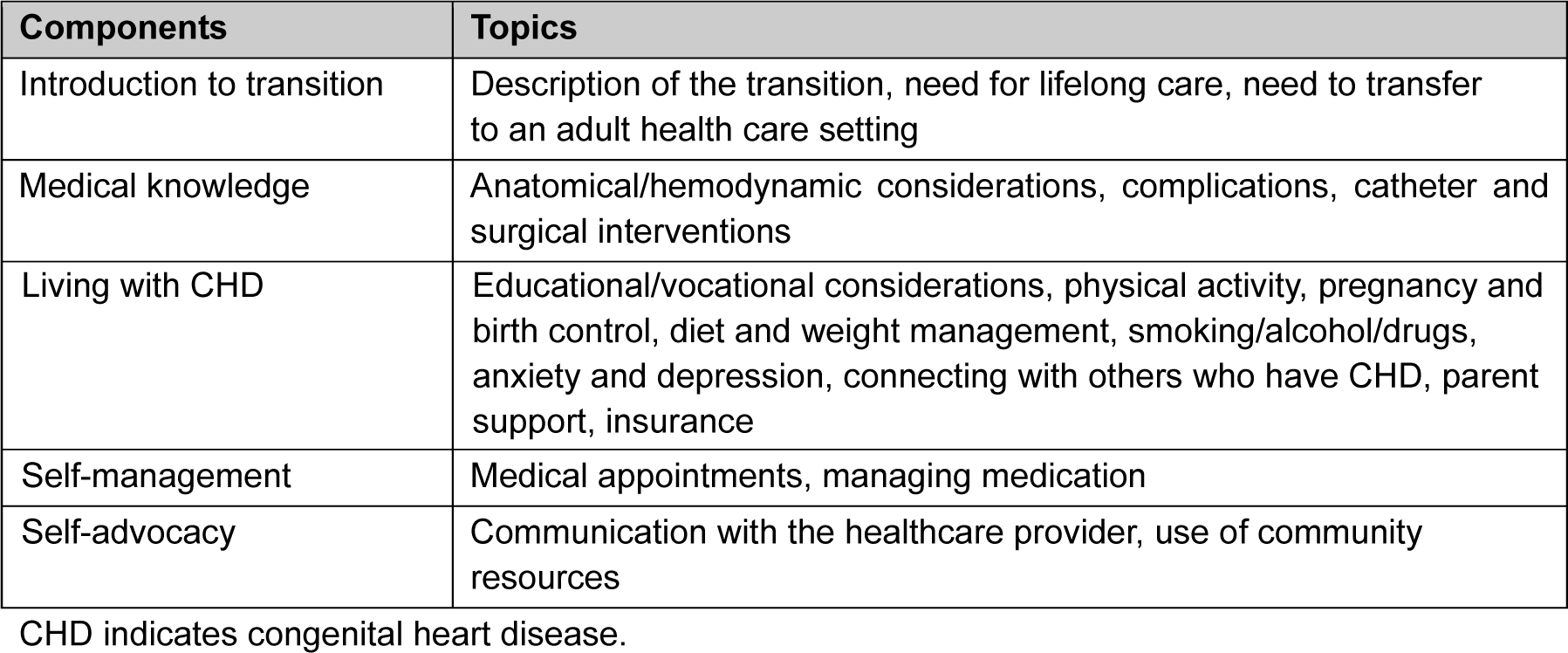
Transition Program Components for Adolescents and Young Adults with Congenital Heart Disease.

| Components | Topics |
| --- | --- |
| Introduction to transition | Description of the transition, need for lifelong care, need to transfer to an adult health care setting |
| Medical knowledge | Anatomical/hemodynamic considerations, complications, catheter and surgical interventions |
| Living with CHD | Educational/vocational considerations, physical activity, pregnancy and birth control, diet and weight management, smoking/alcohol/drugs, anxiety and depression, connecting with others who have CHD, parent support, insurance |
| Self-management | Medical appointments, managing medication |
| Self-advocacy | Communication with the healthcare provider, use of community resources |

CHD indicates congenital heart disease.

#### 2.3. Study Design

The inclusion criteria encompassed randomized controlled trials (RCTs), non-randomized controlled trials (NRCTs), cohort studies, and case-control studies, provided they included a control group. Cohort studies, single-group pretest-posttest studies, cross-sectional studies, and descriptive sudies that did not feature a control group were not considered.

### 3. Data Extraction

Two researchers, BRL and SL, independently carried out literature selection. Following this, one researcher (BRL) extracted the data using a predefined data coding form, while two other researchers independently reviewed the data that had been extracted. The data coding form included information such as the general details of the literature, the study design, the characteristics of the participants and interventions, the sample size, the major outcomes, and the risk of bias for each study. The findings from the study selection and data extraction were shared among the researchers, with any discrepancies being resolved through discussion.

### 4. Quality Evaluation

Quality evaluation was conducted using the Revised Tool to Assess the Risk of Bias in Randomized Trials (RoB 2.0)^11^ and the Risk of Bias Assessment tool for Non-randomized Study (RoBANS).^12^ This evaluation was performed by two researchers, BRL and SL. The interrater reliability of the quality evaluation among the researchers was assessed using Cohen’s kappa statistics. In the event of any discrepancies in the assessments between the researchers, these were resolved through discussion and consensus. The certainty of evidence pertaining to each major outcome was analyzed using the Grading of Recommendations Assessment, Development, and Evaluation (GRADE) approach.

### 5. Statistical Analysis

Effect sizes were analyzed using the meta package in R software version 4.2.1. Due to variation in participant age, as well as the composition and methods of interventions across the selected literature, a random-effects model was used to calculate the pooled effect sizes.^13^ When three or more studies reported the same outcomes, the intervention effect estimates were combined.^14^ The effect size for continuous data was calculated using Hedges’ g to account for potential overestimation due to small sample sizes, while odds ratios (ORs) were used for dichotomous data. Additionally, 95% confidence intervals (CIs) for effect sizes were computed.^14^ Heterogeneity was assessed using Cochran’s Q test,^16^ and the extent of heterogeneity was determined using Higgins I^2^ statistic.^13^ Subgroup analyses were performed to investigate whether the timing of effect measurement contributed to heterogeneity and to examine potential differences in loss to follow-up observations based on parental involvement. Publication bias was assessed using funnel plots, the Egger test, and trim-and-fill analysis.^13,17,18^

## RESULTS

### 1. Study Selection Results

We conducted a keyword search and reviewed the titles and abstracts of 13,429 papers, excluding any duplicates. From these, we selected 20 studies. However, four of these studies were abstracts for which the full text was not available, two clinical trials did not report their results, and two clinical trials were still ongoing, thus providing insufficient information about the characteristics or outcomes of transition programs. Additionally, two papers were not related to transition programs, and two were descriptive studies, leading to their exclusion from the analysis. This left eight studies in the initial selection. We also performed a manual search, which yielded an additional 11,351 papers. After reviewing the titles and abstracts of these papers, we selected 13 studies. Among these, six had already been chosen in the initial selection, two did not have full text available, two were not related to transition programs, and one was a descriptive study. This resulted in the inclusion of two additional studies. In total, we selected and analyzed 10 studies (Figure 1).

**Figure 1.**
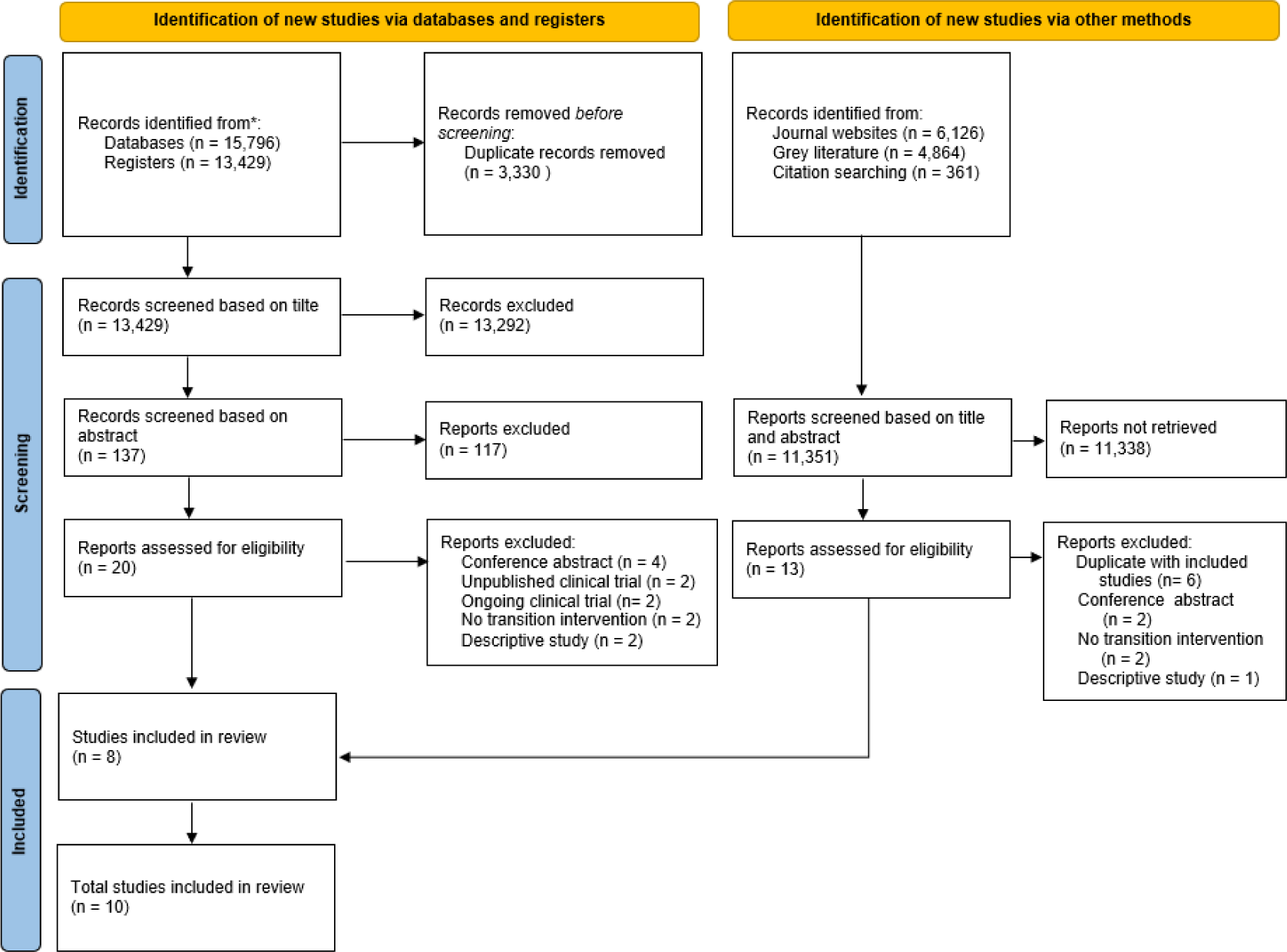
PRISMA flow diagram of the study selection process. PRISMA indicates Preferred Reporting Items for Systematic Reviews and Meta-Analyses.

### 2. General Characteristics of the Study

#### 2.1. Study publication year and nations

One study was published in 2014,^19^ one in 2015,^20^ one in 2017,^21^ two in 2018,^22,23^ one in 2019,^24^ one in 2020,^25^ one in 2021,^26^ and two in 2022.^27,28^ Three studies each were performed in the USA,^22,25,26^ Canada,^19,23,28^ and South Korea,^21,24,27^ while one study was conducted in Belgium^20^ (Table 2).

**Table 2.**
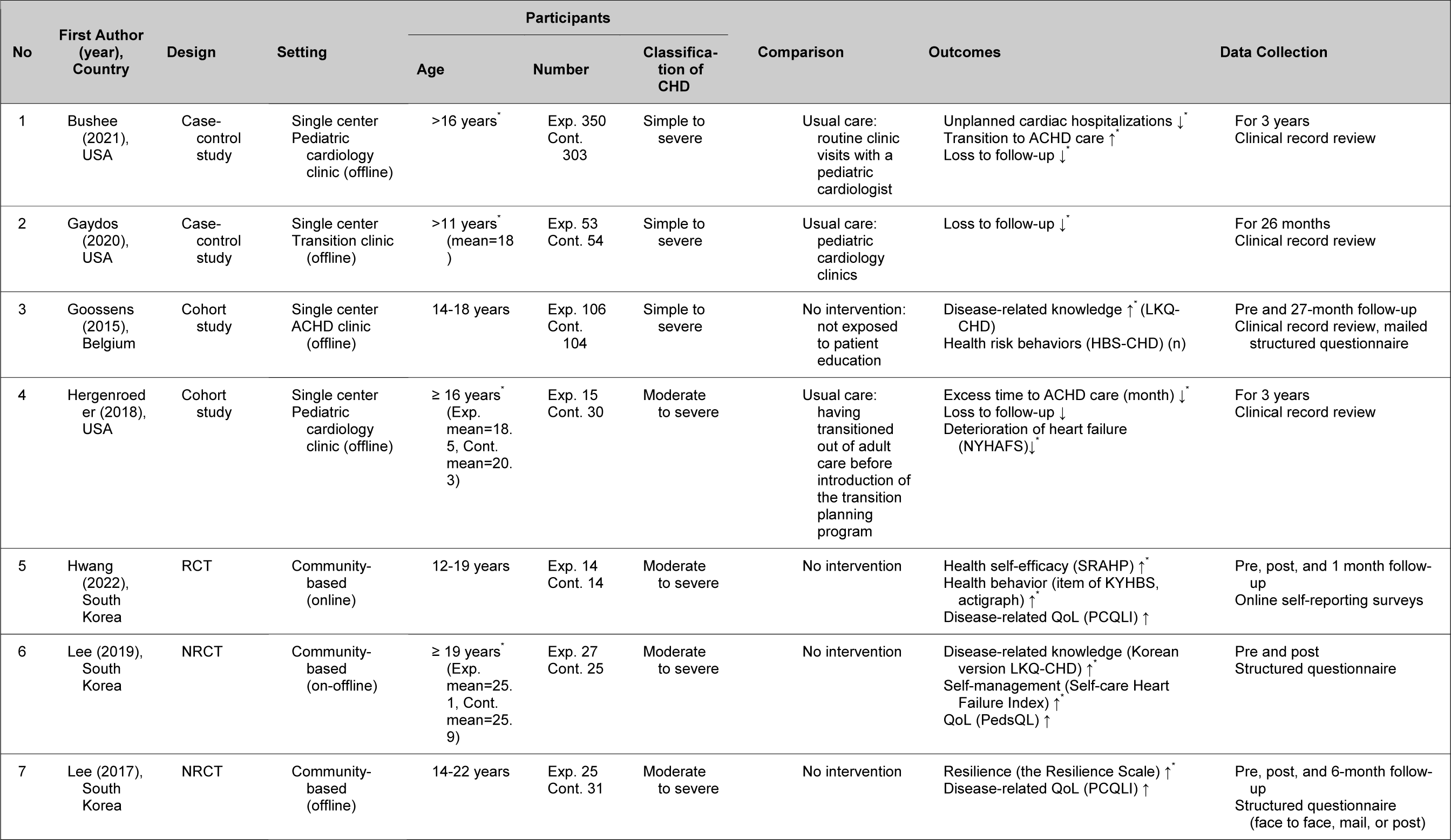
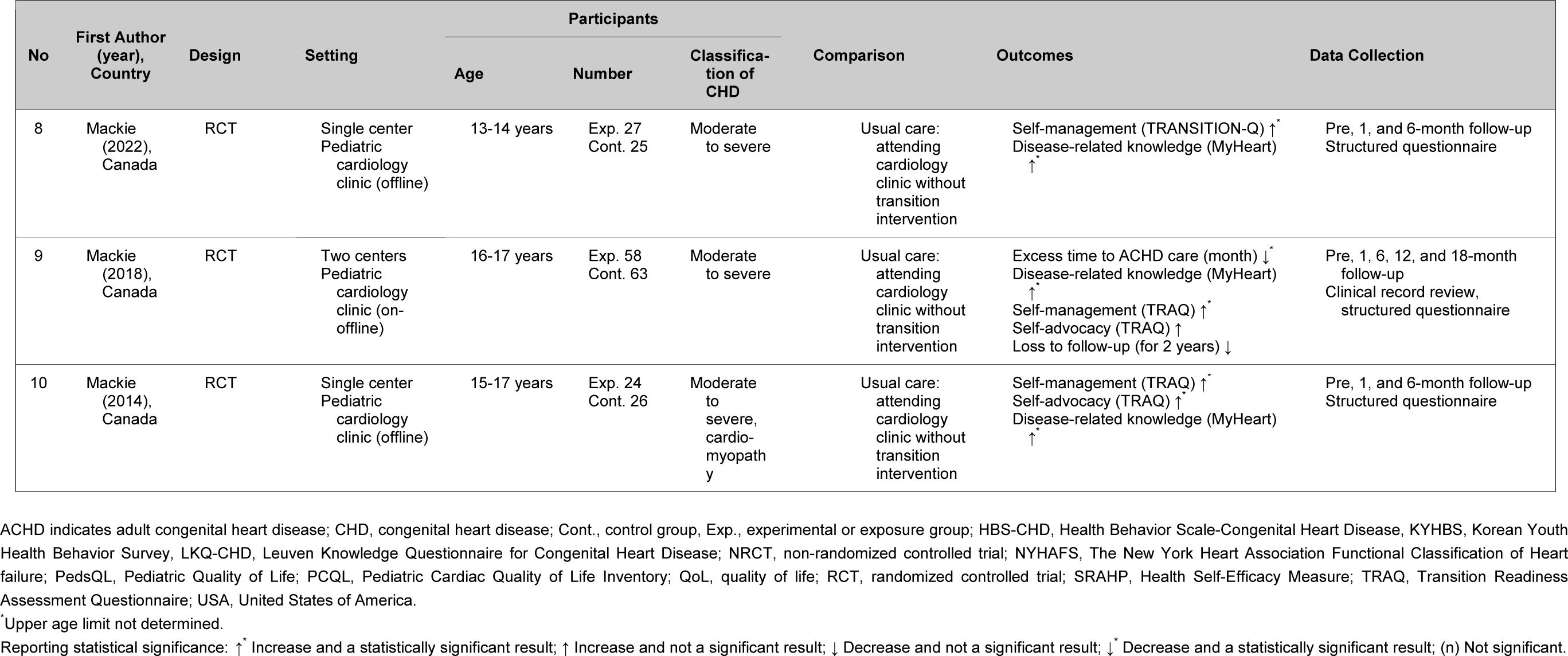
General Characteristics of the Included Studies.

#### 2.2. Study Design

The study designs included four RCTs,^19,23,27,28^ two NRCTs,^21,24^ two cohort studies,^20,22^ and two case-control studies^25,26^ (Table 2).

#### 2.3. Participants

The samples in the 10 studies totaled 1,297 individuals, with a mean age ranging from 13 to 25 years (i.e., adolescents and young adults). If individuals aged 18 and above are classified as young adults,^6^ there were three studies targeting adolescents,^19,23,28^ one study targeting young,^24^ and six studies that broadly encompassed both adolescents and young adults (Table 2).^20–22,25–27^

Regarding the complexity of CHD, seven studies included individuals with moderate and severe complexity CHD,^19,21–24,27,28^ with one of them also incorporating cardiomyopathy.^19^ Three studies encompassed individuals with simple, moderate, and severe complexity CHD (Table 2).^20,25,26^

#### 2.4. Outcomes

In the 10 studies, outcomes were reported for disease-related knowledge,^19,20,23,24,28^ loss to follow-up,^22,23,25,26^ self-management,^19,23,24,28^ quality of life (QoL),^21,24,27^ excess time between pediatric and adult congenital heart disease (ACHD) care,^22,23^ self-advocacy,^19,23^ health and health risk behavior,^20,27^ transition to ACHD care,^26^ unplanned cardiac hospitalizations,^26^ and deterioration of heart failure status^22^ (Table 2). Among these, disease-related knowledge was the most frequently measured outcome, assessed in 5 studies. Conversely, outcomes related to the impact on participants’ health, such as unplanned cardiac hospitalizations and deterioration of heart failure status, were each evaluated in only one study. Detailed outcome statistics for each specific study can be found in Table S1.

### 3. Intervention Characteristics

The characteristics of interventions are summarized and presented in Table 3. Among the included transition programs, six were not based on any specific model or theory. One study utilized guidelines and tools from GotTransition© (https://www.gottransition.org/),^25^ two were based on Bandura’s^29^ self-efficacy theory,^24,27^ and one applied the five psychosocial factors of resilience.^21^

The frequency of interventions varied, with four studies offering interventions once,^19–21,28^ one study providing two interventions,^23^ one study conducting interventions weekly for 4 weeks,^27^ and one study offering interventions weekly for 6 weeks.^24^ Two studies had varying intervention frequencies, with one providing interventions annually until the transfer to ACHD care was complete,^26^ and one providing interventions at least every 6 months until completion of the transfer.^25^ One study did not report the exact frequency of interventions.^22^

**Table 3.** Characteristics of the Interventions of the Included Studies.

| First Author<br>(year),<br>country | Model or<br>Theory | Frequency<br>(duration) | Type | Key Components |  |  |  |  | Provider | Participation<br>of Parents | Other Characteristics |
| --- | --- | --- | --- | --- | --- | --- | --- | --- | --- | --- | --- |
|  |  |  |  | Introduction<br>to Transition | Medical<br>Knowledge | Living with<br>CHD | Self-Management | Self-Advocacy |  |  |  |
| Bushee<br>(2021),<br>USA | - | 1 session or<br>more, yearly<br>(NR) | IP | b,c | d,e,f | o | p,q | r | Transition team:<br>ACHD nurses,<br>physicians,<br>and a social<br>worker | Family included | - |
| Gaydos<br>(2020),<br>USA | GotTransi-<br>tion®<br>guideline | 1 session or<br>more, minimum<br>6 months<br>interval (NR) | IP | a,c | d,e | h,i,m | p,q | s | Transition team:<br>pediatric<br>cardiologist<br>and ACHD NP | Family included | Used an electronic<br>registry to track<br>patients' progress<br>and continuation of<br>care |
| Goossens<br>(2015),<br>Belgium | - | 1 session (15-30<br>min) | IP | c | d,e,f | g,h,i,j,k | p,q | r | ACHD APN team | Not included | Used a computerized<br>checklist in<br>education |
| Hergenroeder<br>(2018),<br>USA | - | NR | IP | a,c | d | o | p,q | r | Two research<br>nurses | Not included | Used an EMR-based<br>transition planning<br>tool to navigate<br>patient's transition |
| Mackie<br>(2022),<br>Canada | - | 1 session (1 hr) | IP | a | d,e,f | i | p,q | r,s | Two nurses | Not included | Used a MyHealth<br>passport for the<br>creation of a<br>portable health<br>summary |
| Mackie<br>(2018),<br>Canada | - | 2 sessions,<br>2-month<br>interval<br>(1-1.5<br>hr/session) | IP | a | d,e,f | i,k | p,q | r,s | Two nurses | Not included | Used a MyHealth<br>passport for the<br>creation of a<br>portable health<br>summary |
| Mackie<br>(2014),<br>Canada | - | 1 session (1 hr) | IP | a | d,e,f | i,k | p,q | r,s | Cardiology<br>nurses | Not included | Used a MyHealth<br>passport for the<br>creation of a<br>portable health<br>summary |
| Hwang<br>(2022),<br>South<br>Korea | Self-efficacy<br>theory | 4 sessions,<br>1 session/week<br>(60<br>min/session) | GP | a,b | d,f | g,h,j,l,m,n | q | r | Nurse | Not included | Performed self-management and<br>1:1 coaching |

**Table 3. Continued**
| First Author<br>(year),<br>country | Model or<br>Theory | Frequency<br>(duration) | Type | Key Components |  |  |  |  | Provider | Participation<br>of Parents | Other Characteristics |
| --- | --- | --- | --- | --- | --- | --- | --- | --- | --- | --- | --- |
|  |  |  |  | Introduction<br>to Transition | Medical<br>Knowledge | Living with<br>CHD | Self-Management | Self-Advocacy |  |  |  |
| Lee (2019),<br>South<br>Korea | Self-efficacy<br>theory | 6 sessions,<br>1 session/week<br>(10-90 min<br>/session) | GP | b | d,e | h,i,j,l,m | p,q | r | Nurses | Not included | Performed self-management and<br>1:1 coaching |
| Lee (2017),<br>South<br>Korea | Resilienc<br>e<br>(psych<br>o-<br>social<br>factor) | 1 session (300<br>min) | GP | b | d,f | g,h,i,j,k,m | p,q | r | Doctors, nurses,<br>NPs,<br>researchers | Not included | Included a group<br>exercise program |

ACHD indicates adult congenital heart disease; APN, advanced practice nursing; CHD, congenital heart disease; GP, group program; IP, individual program; NP, nurse practitioner; NR, not reported; USA, United States of America.
<sup>a</sup> Description of the transition.
<sup>b</sup> Need for lifelong care.
<sup>c</sup> Need to transfer to adult health care setting.
<sup>e</sup> Complications.
<sup>f</sup> Catheter and surgical interventions.
<sup>h</sup> Physical activity.
<sup>i</sup> Pregnancy and birth control.
<sup>j</sup> Diet and weight management.

Seven studies provided individualized transition programs tailored to each patient’s condition and needs,^19,20,22,23,25,26,28^ while three provided group transition programs that involved group activities such as sharing experiences and exercising together among individuals with CHD.^21,24,27^ Among the group transition programs, two involved creating self-management plans and providing one-on-one coaching from nurses via phone or email during the self-management process.^24,27^

The topics covered by almost all (9 or 10) of the transition programs comprised anatomical-hemodynamic considerations (n=10), managing medications (n=10), medical appointments (n=9), and communication with the healthcare provider (n=9). In contrast, lifestyle topics, such as educational/vocational considerations (n=3), anxiety and depression (n=3), insurance (n=2), and parent support (n=1), were covered in three or fewer transition programs.

Nurses were the primary providers of the interventions in seven studies,^19,20,22–24,27,28^ while in two studies, the intervention was delivered by both nurses and physicians.^21,25^ Additionally, one study involved a multidisciplinary team,^26^ and in all cases, nurses were part of the intervention delivery. Two studies included parents in the transition program, considering their opinions when determining the timing of transferring to an adult care environment or providing brochures on transferring health management responsibilities to their children.^25,26^

Three studies used electronic tools to efficiently manage participants during the transition process. Gaydos et al.^25^ created an electronic registry to manage the progress of the transition program and follow-up. Goossens et al.^20^ used a computerized checklist during education to differentiate information that had been discussed with participants or that participants were already aware of. Furthermore, Hergenroeder et al.^22^ developed an electronic medical record (EMR)-based transition planning tool to track and assess participants, which served as the basis for providing the transition program.

### 4. Risk of Bias Assessment

Out of the 10 selected studies, four RCTs^19,23,27,28^ were evaluated using RoB 2.0 (5 domains), while the remaining six studies^20–22,24–26^ were assessed using RoBANS (6 domains). The Cohen’s kappa for quality assessment across a total of 56 domains was 0.75 (95% CI=0.61-0.85), indicating moderate agreement.^30^

#### 4.1. RCTs

(1) Mackie et al.^19^ employed random allocation, clustering by the week of attendance in the cardiology clinic. However, a detailed description of the allocation sequence was not provided, resulting in a rating of “some concerns.” (2-4) In all four RCTs,^19,23,27,28^ there were no deviations from the intended interventions, and the risk of bias in terms of missing data and outcome measurement was low. (5) Two studies by Mackie et al.^23,28^ had follow-up durations ranging from 6 to 18 months, and some mean and standard deviation (SD) values were not reported. This raised concerns about potential bias. (6) The overall bias assessment indicated that one study^27^ had a low risk of bias, while three studies^19,23,28^ were rated as having some concerns regarding the risk of bias (Figure 2).

**Figure 2.**
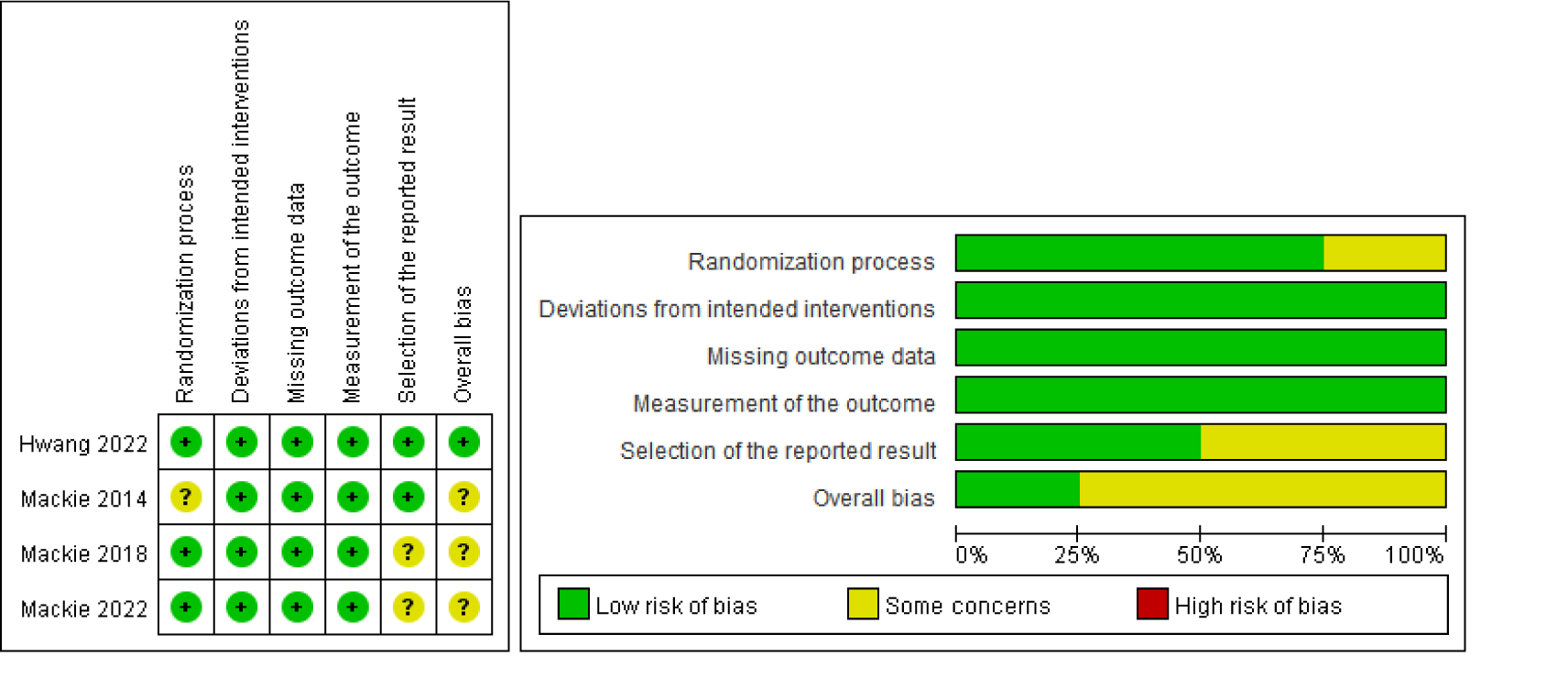
RoB 2.0 for randomized controlled trials. ROB indicates risk of bias assessment using a revised tool to assess the risk of bias in randomized trials.

#### 4.2. NRCTs, Cohort Studies, and Case-Control Studies

(1) The cohort studies conducted by Goossens et al.^20^ and Hergenroeder et al.^22^ did not report baseline differences in outcome variables or utilized a historical control group, resulting in a high risk of bias in the selection of participants. (2) Bias due to confounding variables was low in all six studies^20–22,24–26^. (3) Two studies^21,24^ collected all outcomes using self-reported tools, which raised concerns about the risk of bias. (4) Lee and Jung’s study^24^ did not provide details about blinding of outcome assessors, leading to an unclear risk of bias. (5) The study of Goossens et al.^20^ reported detailed results only for some participants who participated in all outcome measurements within the control group, resulting in an unclear risk of bias. (6) The study of Goossens et al.^20^ did not report the mean and SD for secondary outcomes, leading to a high risk of bias (Figure 3).

**Figure 3.**
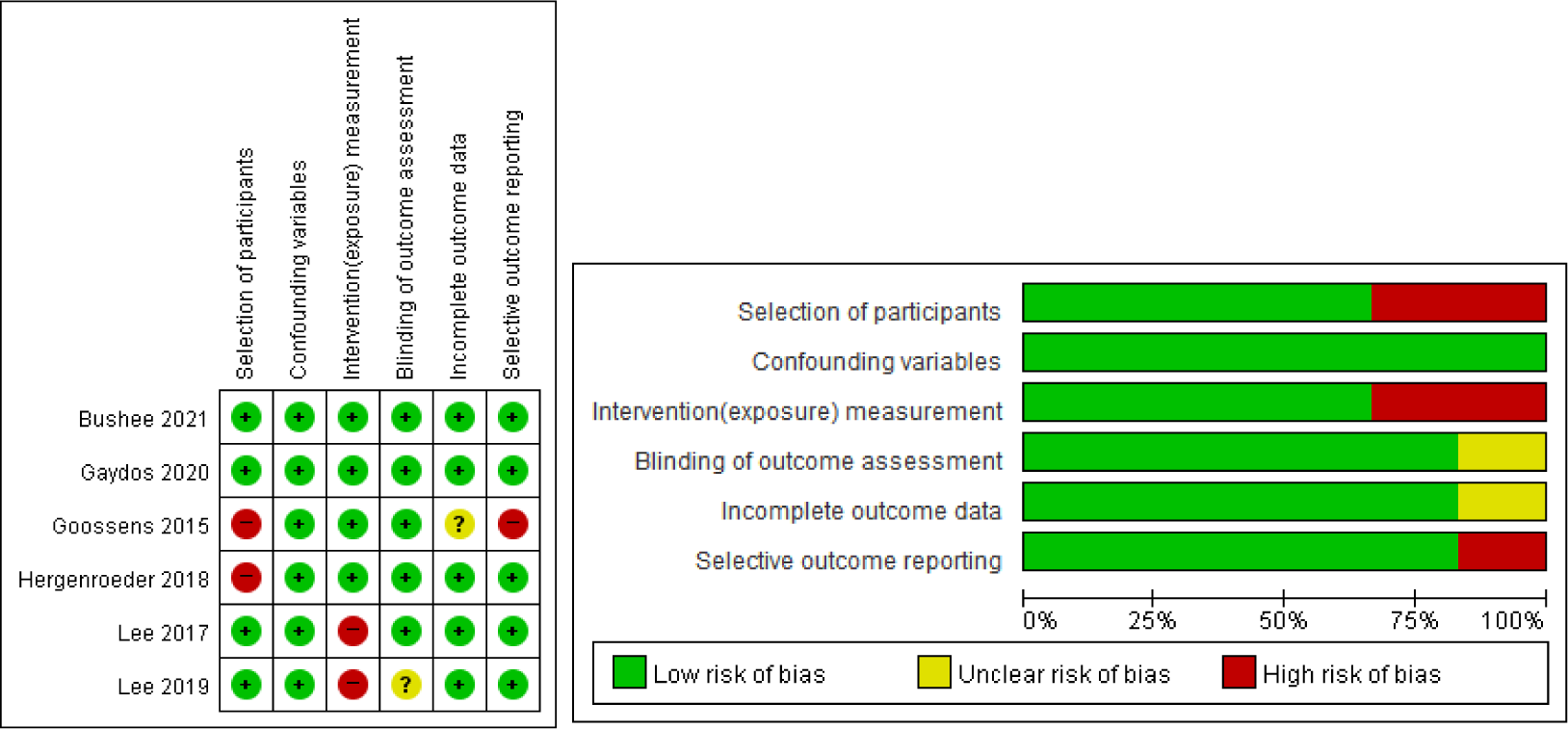
Risk of bias assessment using RoBANS. RoBANS indicates the risk of bias assessment tool for non-randomized studies.

### 5. Results of the Meta-Analysis

#### 5.1. Disease-related Knowledge

In the five studies reporting disease-related knowledge, Hedge’s g was 0.89 (95% CI=0.29-1.48), indicating a large effect size that was statistically significant. Heterogeneity testing revealed a Cochran’s Q value of 32.7 (*p*<.001, df=4), indicating heterogeneity in effect sizes across studies, and the degree of heterogeneity was substantial (Higgins I^2^=87.8%; Figure 4). Since the timing of outcome measurements varied among the included studies, a subgroup analysis was conducted by dividing studies into those reporting immediate post-intervention results and those reporting results from follow-up assessments (Figure 5). Disease-related knowledge showed a significant increase in the experimental group compared to the control group when measured immediately after the intervention (2.26, 95% CI=1.57-2.95). Furthermore, in follow-up assessments of 6 months or more, the experimental group exhibited a significant increase in disease-related knowledge compared to the control group (0.55, 95% CI=0.18-0.93), and this difference in effect sizes between the two groups was also significant (F^2^=18.07, df=1, *p*<.001).

**Figure 4.**
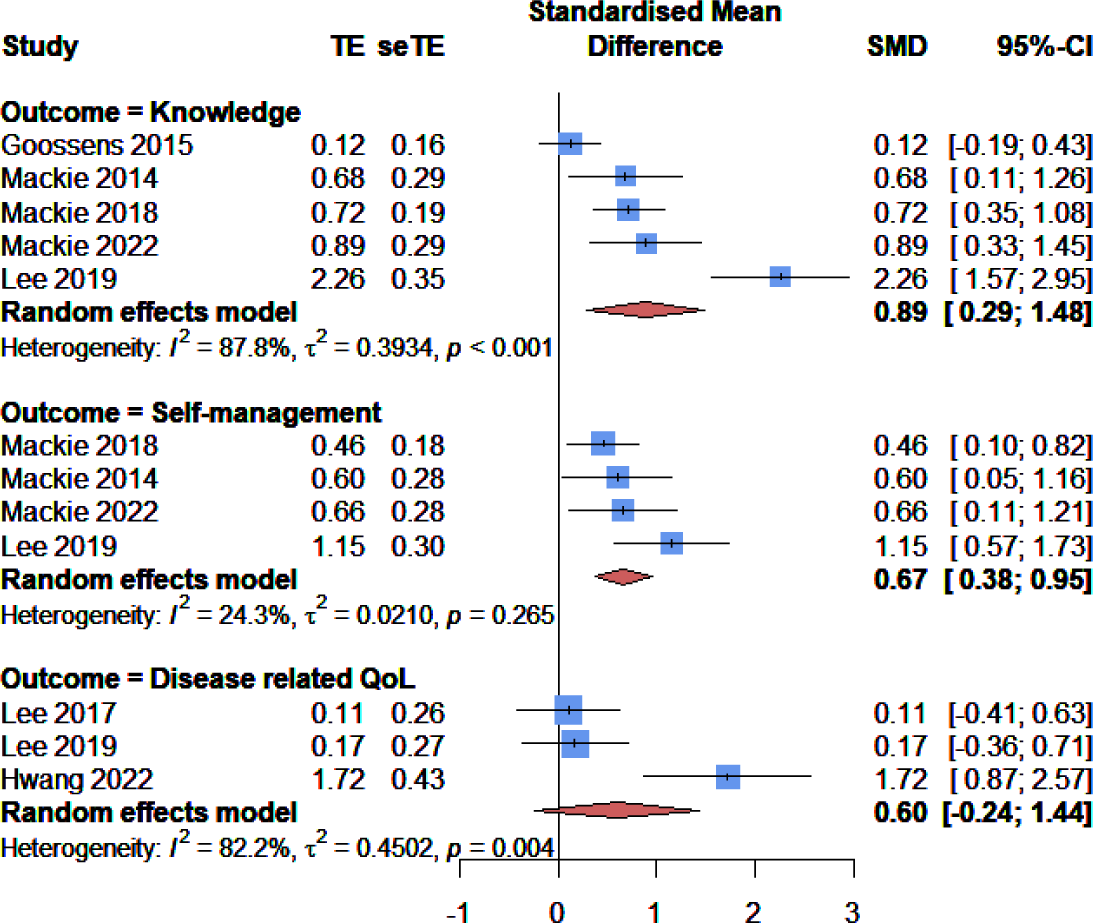
Forest plot of the effects of transition programs for adolescents and young adults. QoL indicates quality of life.

**Figure 5.**
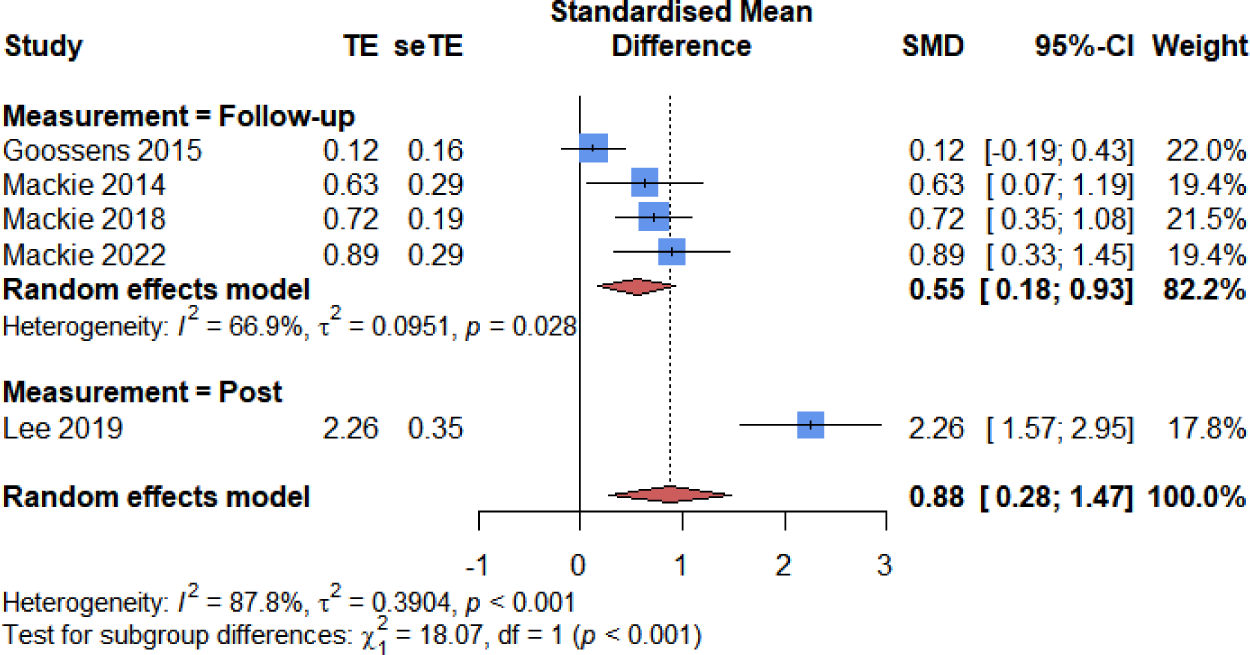
Forest plot of the disease-related knowledge effect of transition programs for adolescents and young adults according to the timing of measurement.

#### 5.2. Self-management

In the four studies reporting self-management, Hedge’s g was 0.67 (95% CI=0.38-0.95), indicating a moderate effect size that was statistically significant. Heterogeneity testing showed a Cochran’s Q value of 3.97 (*p*=.265, df=3), suggesting that the effect sizes across studies were not heterogeneous, and the degree of heterogeneity was low (Higgins I^2^=24.3%; Figure 4).

#### 5.3. Disease-related QoL

In the three studies reporting disease-related QoL, Hedge’s g was 0.60 (95% CI=-0.24-1.44), indicating a moderate effect size that was not statistically significant. Heterogeneity testing showed a Cochran’s Q value of 11.22 (*p*=.004, df=2), indicating heterogeneity in the effect sizes across studies, and the degree of heterogeneity was substantial (Higgins I^2^=82.2%; Figure 4). However, since the number of studies included in the effect size synthesis was less than four, a moderation analysis could not be conducted.^14^

#### 5.4. Loss to Follow-Up

The pooled effect size for loss to follow-up, reported in four studies, showed that the experimental group receiving the transition program had a 0.41-fold (95% CI=0.22-0.77) lower rate of loss to follow-up compared to the control group, and this difference was statistically significant. The converted value for this effect size, represented by Hedge’s g, was 0.49 (95% CI= 0.15-0.83), indicating a moderate effect size. Heterogeneity testing revealed a Cochran’s Q value of 4.88 (*p*=.181, df=3), indicating no heterogeneity in effect sizes across studies, and the degree of heterogeneity was moderate (Higgins I^2^=38.5%; Figure 6). In line with the recommendation to include parents in transition programs to facilitate the transfer of self-management responsibilities,^6,31^ we analyzed differences in the effect of loss to follow-up according to parental involvement. The results showed that when parents participated in the transition program, the experimental group had a 0.36-fold (95% CI=0.22-0.59) reduction in the loss to follow-up rate compared to the control group, and this was statistically significant. However, when parents did not participate, the OR for loss to follow-up was not statistically significant, and the difference in effect size between the two groups was also not significant (χ^2^=0.01, df=1, *p*=.936; Figure 7).

**Figure 6.**
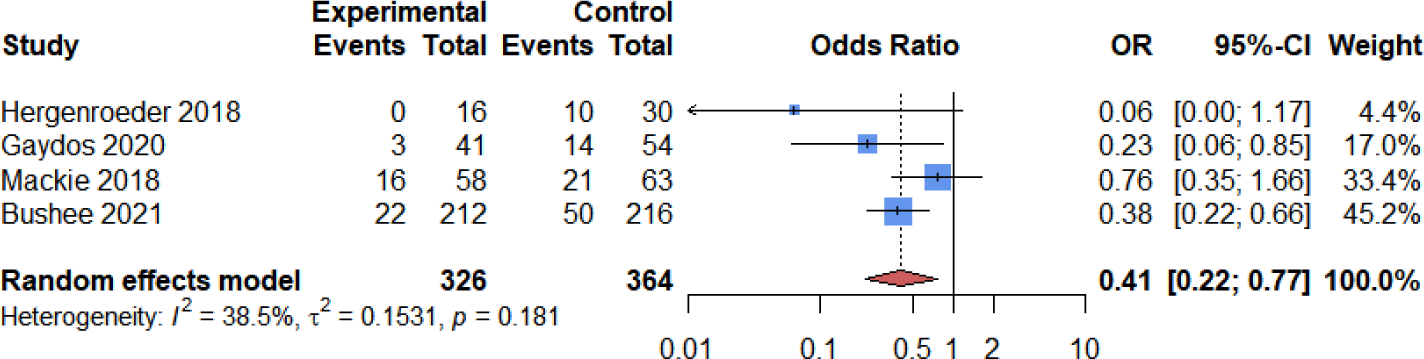
Forest plot of the effect of transition programs for adolescents and young adults on loss to follow-up.

**Figure 7.**
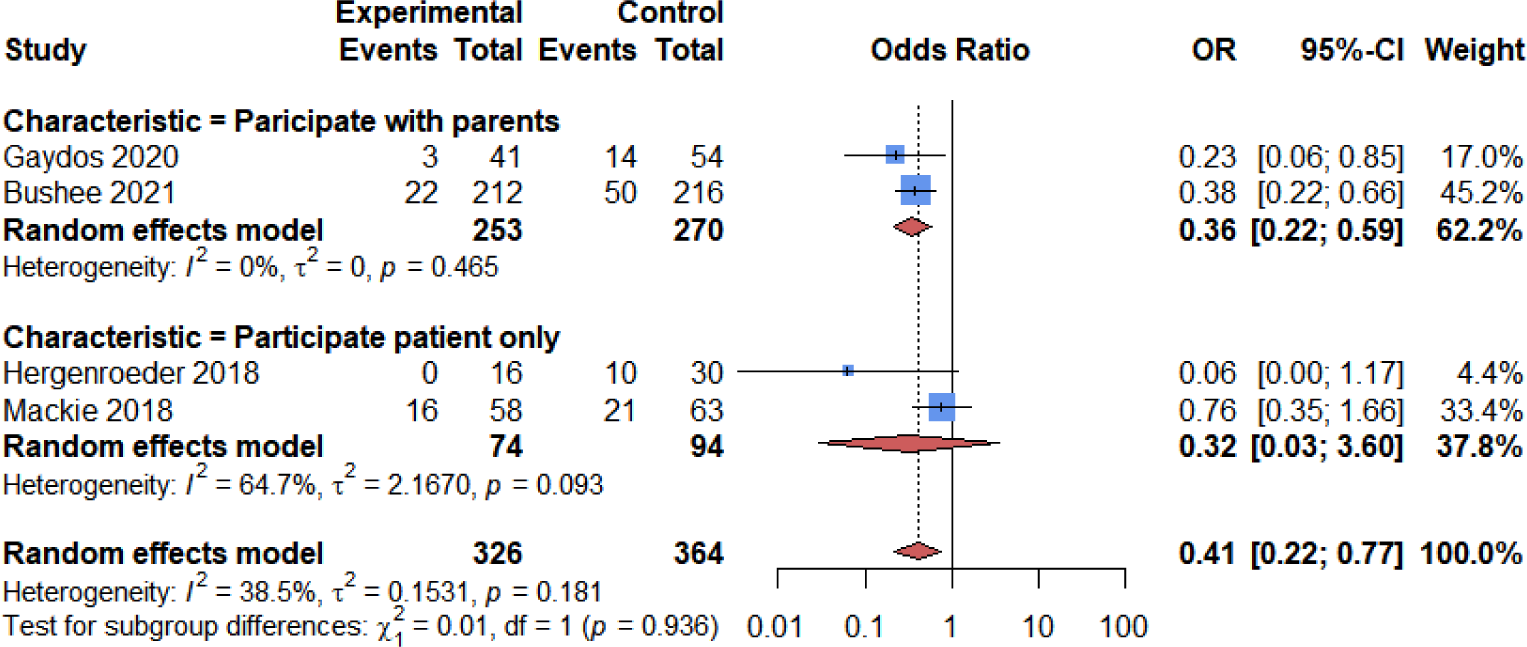
Forest plot of the effect of transition programs for adolescents and young adults on loss to follow-up according to parents’ participation.

### 6. Publication Bias

Publication bias was assessed using the effect size of the primary outcome for each study. A right-skewed funnel plot was observed, and the Egger test indicated that the regression equation was not statistically significant (t=2.20, *p*=.059). The Hedge’s g, adjust ed using the trim and fill method, decreased compared to the unadjusted Hedge’s g. However, both effect sizes remained statistically significant. It was concluded that even though publication bias was present, it did not have a significant impact on the results of the studies (Figure S1; Figure S2).

### 7. Quality of Evidence

The quality of evidence for the outcome variables included in the meta-analysis—namely, disease-related knowledge, loss to follow-up, self-management, and disease-related QoL—was assessed. Disease-related knowledge and loss to follow-up were evaluated as having low-quality evidence, indicating limited reliability of effect estimates. Self-management and disease-related QoL were assessed as having very low-quality evidence, suggesting that there is almost no certainty regarding the effect estimates. Detailed results are provided in Table S2.

## DISCUSSION

### 1. Research Characteristics

Transition programs for CHD patients have been implemented in the USA, Canada, South Korea, and Belgium since 2014. These programs primarily target adolescents and young adults, with an average age range of 13-25 years. They encompass a broad age spectrum, from adolescents to young adults (n=6), with the majority focusing on those with moderate to severe complexity CHD (n=7). An examination of these transition programs reveals that the selected studies generally align with the recommendations from the American Heart Association’s scientific statement^5^ and the global consensus statement^6^ from various national heart associations worldwide. However, there are differences in the specific topics included in the major components of the transition programs, the frequency and duration of interventions, parental involvement, outcomes, and tools. Thus, there was significant heterogeneity among the studies.

Among the 10 transition programs reviewed, over nine studies incorporated topics related to the anatomical-hemodynamic considerations of CHD (n=10), medication management (n=10), medical appointments (n=9), and communication with healthcare providers (n=9). The anatomical-hemodynamic considerations of CHD are part of the medical knowledge component of the transition program, while medication management and medical appointments fall under the self-management component. Communication with healthcare providers, in contrast, is part of the self-advocacy component.^5^ In other words, transition programs for adolescents and young adults with CHD focus on enhancing their independent self-management and self-advocacy skills, grounded in a thorough understanding of the disease. Experts in the field of transition concur that acquiring skills related to medical knowledge, self-management, and self-advocacy is vital among the various topics covered in transition programs.^31–33^ Self-management and self-advocacy skills can bolster a CHD patient’s medical independence, decision-making capacity in healthcare, and control over their psychosocial environment.^31^ A lack of medical knowledge can hinder patients from developing self-management and self-advocacy skills.^34^ Therefore, it is crucial for transition programs to incorporate education and role supplementation concerning medical knowledge to understand the condition, self-management skills for managing the condition, and self-advocacy skills to enhance decision-making and communication.

Interventions related to educational/vocational considerations (n=3), anxiety and depression (n=3), and parent support (n=1) were relatively underrepresented in transition programs for lifestyle management. Education for patients with CHD is closely linked to adult employment and income.^5^ Adolescents and young adults with CHD often encounter educational delays and face difficulties in making career choices.^31,35^ They are less likely to pursue higher education or vocational training compared to their peers without CHD.^36^ Therefore, transition programs for adolescents and young adults with CHD should consider incorporating additional educational and vocational interventions. Moreover, individuals with CHD have a higher lifetime prevalence of anxiety and depression compared to the general population,^37^ especially during the adolescent and young adult years when the transition process involves shifting disease management responsibilities from parents to the individual. This shift can intensify psychological distress in these individuals.^35^ Parent support is also recognized as a crucial factor in the psychosocial well-being of CHD patients.^38^ Therefore, when developing transition programs, it is important to include psychological interventions that are based on assessments of anxiety and depression. Additionally, considering parental support during the process of empowering patients to manage their own healthcare decisions can prove beneficial.^5,39^

Among the 10 transition programs analyzed, 7 were individual programs, outnumbering the group programs. This contrasts with the findings of a systematic review of transition programs for pediatric patients with chronic conditions, where individual and group programs were reported to be equally represented.^9^ This discrepancy may be due to the diverse nature of cardiac diseases within CHD, which vary in severity, characteristics, and management strategies based on individual patients.^40^ Consequently, a more personalized approach is often favored. Notably, all three transition programs reported in South Korea were group programs. This could be because in South Korea, ACHD care is not distinct from pediatric care.^41^ Therefore, these programs tend to focus more on enhancing adult disease management in a community-based group setting, rather than primarily aiming to transition to ACHD care.

Among the transition program studies analyzed, four were observational studies, while seven demonstrated a risk of bias. The primary sources of bias included the omission of all outcome values when measurements were taken multiple times, the use of historical control groups, the failure to clearly report baseline differences in outcome variables, and self-reporting. Therefore, it is crucial to consider these biases when examining the effects of transition programs in future research. Additionally, some studies did not provide detailed descriptions of program aspects such as the exact age of participants, program frequency, and duration. Consequently, future studies should adhere to reporting guidelines to ensure that transition programs can be replicated.

### 2. Effects of Transition Interventions

After the transition programs, the participants showed a substantial increase in disease-related knowledge, with a large effect size. In this context, disease-related knowledge refers to understanding medical aspects related to CHD, including its complications, medical interventions, and disease management strategies.^42,43^ Youth with CHD often have limited knowledge about topics such as physical activity, diet, and sexual health related to disease management,^44–46^ and they frequently struggle to accurately locate their specific cardiac abnormalities and comprehend the complexities of cardiac defects and complications.^44,46^ During the transition program, it is recommended to employ strategies such as creating portable electronic files like the MyHealth passport to provide information about heart conditions, and using diagrams to educate patients about anatomical defects.^5^ In the transition programs included in this study, similar approaches were likely employed. These programs likely utilized visual materials, such as MyHealth passports and anatomical diagrams, to educate participants about medical knowledge, contributing to the improvement in disease-related knowledge. Notably, the study conducted by Lee and Jung^24^, which reported the largest effect size, was based on self-efficacy theory. In addition to written educational materials, they produced successful disease management videos to provide audio-visual materials. Unlike other studies that delivered the intervention 1-2 times, they implemented the program over 6 weeks and augmented knowledge comprehension by offering weekly phone and email support to address participants’ queries.

Disease-related knowledge showed a larger effect size immediately after the intervention than in follow-up assessments conducted after the intervention. According to the Forgetting Curve Theory^47^, knowledge tends to be forgotten at a rate of approximately 80% within 1 month. Therefore, it is plausible that the effect size diminished in assessments conducted after a period of 6 months or more. Given that the effectiveness of intervention on disease-related knowledge may wane over time, regular re-evaluations and supplementary education are essential. However, in this meta-analysis, the follow-up periods across the studies varied from 6 to 26 months, highlighting inconsistencies between studies. Further research is required to ascertain the optimal timing for reassessment.

Self-management among participants saw a moderate increase following the transition program. This concept of self-management includes behavioral aspects such as obtaining prescribed medications when necessary, arranging follow-up appointments, and attending medical consultations.^48^ To encourage changes in health behavior, interventions that not only impart knowledge but also promote self-monitoring, risk communication, and social support have proven effective. As such, interventions should incorporate behavior change strategies that combine knowledge, awareness, and facilitation.^49^ The transition programs analyzed in this study utilized various role supplementation techniques for awareness and facilitation. These included discussions, encouragement, sharing and providing experiences, and offering relevant resources. Of particular note is the study by Lee and Jung,^24^ which reported a large effect size. This study provided participants with a self-management diary to encourage self-monitoring of medication adherence and symptoms. They also offered one-on-one telephone counseling to promote self-management, and mailed necessary resources to participants. Additionally, the program facilitated discussions among participants, allowing them to share their self-management experiences.

Loss to follow-up decreased after the transition programs, with a moderate effect size. In a previous meta-analysis by Moons et al.^1^, the effect size for discontinuation of follow-up in CHD patients after transition programs was not statistically significant. However, in this study, the addition of one more study to the effect size pooling conducted by Moons et al.^1^ yielded a significant effect on loss to follow-up. Furthermore, a meta-analysis by García-Rodríguez et al.,^50^ which included various chronic disease populations, also found a significant effect size for clinic drop-out. By combining the results of the previous meta-analysis with the findings of this study, it can be deduced that transition programs may help reduce loss to follow-up. A recent systematic review identified structured transition programs led by nurses and formal handovers to ACHD care as strategies to decrease treatment discontinuation in CHD patients.^51^ The transition programs evaluated for loss to follow-up in this study employed structured transition programs, which included guidelines from Got Transition©, EMR-based transition planning tools, and shared patient information among healthcare providers. These programs were primarily nurse-led or included nurses as part of the transition team. Notably, the study by Mackie et al.,^23^ which reported a significant effect size on the reduction in loss to follow-up, involved pediatric cardiac nurses visiting an ACHD outpatient clinic with patients and directly transitioning them, thereby reducing the rate of loss to follow-up.

The analysis found no statistically significant differences in loss to follow-up based on parental involvement. However, it’s important to note that the number of studies included in this meta-analysis was limited, and the specific roles of parents and the details of their interventions in transition programs were not thoroughly described. Parents can play a significant role in the follow-up process during transitions by engaging in discussions with their children about the follow-up before visiting the outpatient clinic, preparing a list of questions for healthcare providers, and encouraging their children to communicate directly with these providers and ask questions.^6^ When planning future transitions, parents should be involved from the beginning, and they should receive the necessary information to serve as positive resources throughout the transition process. Further analysis is required to evaluate the effectiveness of such transition programs.

The effect of the transition programs on disease-related QoL was not statistically significant. The disease-related QoL in adolescents and adults with CHD is influenced by the complexity of the CHD, clinical symptoms, and complications, with only limited associations with demographic and socio-economic factors.^52–54^ Therefore, to enhance disease-related QoL in individuals with CHD, the focus should be on alleviating clinical symptoms and complications associated with CHD. A prior meta-analysis indicated an improvement in disease-related QoL when individuals with CHD participated in an exercise program lasting from 10 weeks to 3 months, which improved cardiac function and reduced the need for medication.^55^ In this study, the transition programs that evaluated disease-related QoL also incorporated topics aimed at improving cardiac health through lifestyle factors such as physical activity, diet, and weight management. These programs were intended to motivate participants to actively participate in exercise and make dietary modifications. However, these programs were relatively brief, lasting only 1 day or 6 weeks. To effectively improve disease-related QoL, future programs should include topics related to lifestyle management, focusing on enhancing cardiac function and clinical symptoms, and should be implemented over longer periods, such as 10 weeks or more, to evaluate their effectiveness.

This study marks the first attempt to conduct a meta-analysis of the varied effects of transition programs for individuals with CHD, by identifying five key components of CHD-targeted transition programs and analyzing appropriate programs based on these components. By defining transition programs clearly and conducting an exhaustive literature search with heightened sensitivity, this research sought to strengthen the validity of effect assessment. This was achieved by selecting studies that included control groups and analyzing their outcomes to yield meaningful conclusions. Consequently, this study carries significant importance as it provides a current overview of transition programs for adolescents and young adults with CHD. This serves as an evidence base for researchers and practitioners interested in developing and implementing transition programs for CHD patients in the future.

### 3. Limitations

This study has several limitations: First, the specific topics included within the transition program components and the structures of the programs themselves varied, resulting in significant heterogeneity among the studies. However, due to the limited number of studies, it was difficult to determine which factors within the intervention positively impacted specific outcomes. Second, in addition to the heterogeneity found in the transition program components, there was also expected heterogeneity due to the broad range of participant ages. However, six out of the ten included pieces of literature encompassed both adolescents and young adults, which complicated the task of conducting a subgroup analysis based on age groups. Third, there was a dearth of studies reporting actual health-related outcomes, such as emergency room visits, hospitalizations, and disease or complication status, which precluded an analysis of their effects. Fourth, transition program studies that did not have a control group, as well as unpublished abstracts and clinical trials (due to insufficient information), were excluded. Lastly, methodological weaknesses in the included studies (risk of bias), high heterogeneity between studies, and small sample sizes resulted in low or very low levels of certainty in the evidence produced by the meta-analysis. Therefore, these limitations should be taken into account when interpreting the results of the analysis.

## CONCLUSIONS

This study conducted a systematic review of transition programs designed to aid adolescents and young adults with CHD in managing their condition as they transition into adulthood. To evaluate the effectiveness of these programs, a comprehensive search of research literature, clinical trials, and grey literature was undertaken. Ten studies that met the selection criteria were ultimately analyzed. The transition programs for adolescents and young adults with CHD focused on providing education and role supplementation to enhance their medical knowledge to understand the disease, their self-management skills, and their self-advocacy skills to improve decision-making and communication. The impact of these transition programs was evident in the improvement of disease-related knowledge, self-management, and a reduction in loss to follow-up. However, there was no significant effect on improving disease-related QoL. It is important to acknowledge that the studies included in this review had methodological weaknesses (risk of bias), a high degree of heterogeneity, and limited sample sizes. As a result, the evidence supporting the effect estimates was of low or very low certainty. Therefore, caution should be exercised when interpreting these results, and further replication studies are recommended for the future.

## Sources of Funding

None.

## Disclosures

This article was adapted from a thesis by Bo Ryeong Lee in partial fulfillment of the requirements for the dissertation at Daegu Catholic University of Korea.

## Supplementary Material

Tables S1-S2

Figures S1-S2

## Data Availability

The datasets are available from the corresponding author on reasonable request.

## Nonstandard Abbreviations and Acronyms

ACHD: adult congenital heart disease
QoL: quality of life

## Notes

### Competing Interest Statement

The authors have declared no competing interest.

### Clinical Trial

CRD42023399026

### Author Declarations

This study was a literature review of previously published studies and was therefore exempt from Institutional Review Board approval.

## REFERENCES

1. Gilboa SM, Devine OJ, Kucik JE, Oster ME, Riehle-Colarusso T, Nembhard WN, Xu P, Correa A, Jenkins K, Marelli AJ. Congenital heart defects in the United States: estimating the magnitude of the affected population in 2010. Circulation. 2016;134(2):101–109. DOI: 10.1161/CIRCULATIONAHA.115.019307.

2. Moons P, Skogby S, Bratt EL, Zühlke L, Marelli A, Goossens E. Discontinuity of cardiac follow-up in young people with congenital heart disease transitioning to adulthood: a systematic review and meta-analysis. J Am Heart Assoc. 2021;10(6):e019552. DOI: 10.1161/JAHA.120.019552.

3. Müller MJ, Norozi K, Caroline J, Sedlak N, Bock J, Paul T, Geyer S, Dellas C. Morbidity and mortality in adults with congenital heart defects in the third and fourth life decade. Clin Res Cardiol. 2022;111(8):900–911. DOI: 10.1007/s00392-022-01989-1.

4. Wray J, Frigiola A, Bull C; Adult Congenital Heart disease Research Network (ACoRN). Loss to specialist follow-up in congenital heart disease; out of sight, out of mind. Heart. 2013;99(7):485–490. DOI: 10.1136/heartjnl-2012-302831.

5. John AS, Jackson JL, Moons P, Uzark K, Mackie AS, Timmins S, Lopez KN, Kovacs AH, Gurvitz M; American Heart Association Adults with congenital Heart Disease Committee of the Council on Lifelong congenital Heart Disease and Heart Health in the Young and the Council on Clinical Cariology, et al. Advances in managing transition to adulthood for adolescents with congenital heart disease: a practical approach to transition program design: a scientific statement from the American Heart Association. J Am Heart Assoc. 2022;11(7):e025278. DOI: 10.1161/JAHA.122.025278.

6. Moons P, Bratt EL, De Backer J, Goossens E, Hornung T, Tutarel O, Zühlke L, Araujo JJ, Callus E, Gabriel H, et al. Transition to adulthood and transfer to adult care of adolescents with congenital heart disease: a global consensus statement of the ESC Association of Cardiovascular Nursing and Allied Professions (ACNAP), the ESC Working Group on Adult Congenital Heart Disease (WG ACHD), the Association for European Paediatric and Congenital Cardiology (AEPC), the Pan-African Society of Cardiology (PASCAR), the Asia-Pacific Pediatric Cardiac Society (APPCS), the Inter-American Society of Cardiology (IASC), the Cardiac Society of Australia and New Zealand (CSANZ), the International Society for Adult Congenital Heart Disease (ISACHD), the World Heart Federation (WHF), the European Congenital Heart Disease Organisation (ECHDO), and the Global Alliance for Rheumatic and Congenital Hearts (Global ARCH). Eur Heart J. 2021;42(41):4213–4223. DOI: 10.1093/eurheartj/ehab388.

7. Carrizosa J, An I, Appleton R, Camfield P, Von Moers A. Models for transition clinics. Epilepsia, 2014;55(Suppl 3):46–51. DOI: 10.1111/epi.12716.

8. Campbell F, Biggs K, Aldiss SK, O’Neill PM, Clowes M, McDonagh J, While A, Gibson F. Transition of care for adolescents from paediatric services to adult health services. Cochrane Database Syst Rev. 2016;4:CD009794. DOI: 10.1002/14651858.CD009794.pub2.

9. Wakimizu R, Sasaki K, Yoshimoto M, Miyazaki A, Saito Y. Multidisciplinary approach for adult patients with childhood-onset chronic disease focusing on promoting pediatric to adult healthcare transition interventions: an updated systematic review. Front Pediatr. 2022;10:919865. DOI: 10.3389/fped.2022.919865.

10. Bidwell S. Database selection and search strategy optimization: the COSI search protocol [abstract]. In: 16th Annual Meeting of the International Society of Technology Assessment in Health Care; 2000 June 19-21; The Hague.

11. Sterne JA, Savović J, Page MJ, Elbers RG, Blencowe NS, Boutron I, Cates CJ, Cheng HY, Corbett MS, Eldridge SM, et al. RoB 2: a revised tool for assessing risk of bias in randomized trials. BMJ. 2019;366:l4898. DOI: 10.1136/bmj.l4898.

12. Kim SY, Park JE, Lee YJ, Seo HJ, Sheen SS, Hahn S, Jang BH, Son HJ. Testing a tool for assessing the risk of bias for nonrandomized studies showed moderate reliability and promising validity. J Clin Epidemiol. 2013;66(4):408–414. DOI: 10.1016/j.jclinepi.2012.09.016.

13. Higgins JP, Thomas J, Chandler J, Cumpston M, Li T, Page MJ, Welch VA (editors). Cochrane Handbook for Systematic Reviews of Interventions version 6.3 (updated February 2022). Cochrane, 2022. Available from www.training.cochrane.org/handbook. Accessed April 20, 2023.

14. Borenstein M, Hedges LV, Higgins JP, Rothestein HR. Introduction to meta-analysis. Chichester, West Sussex: John Wiley & Sons; 2009.

15. Shenhav L, Heller R, Benjamini Y. Quantifying replicability in systematic reviews: the r-value. arXiv. 2015;1502.00088:1-21. Available at: https://arxiv.org/pdf/1502.00088.pdf. Accessed April 30, 2023.

16. Cochran WG. Some methods for strengthening the common χ^2^ tests. Biometrics. 1954;10(4):417–451. DOI: 10.2307/3001616.

17. Duval S, Tweedie R. Trim and fill: a simple funnel-plot-based method of testing and adjusting for publication bias in meta-analysis. Biometrics. 2000;56(2):455–463. DOI: 10.1111/j.0006-341x.2000.00455.x.

18. Harrer M, Cuijpers P, Furukawa TA, Ebert DD. Doing meta-analysis with R: a hands-on guide. Boca Raton, FL and London: Chapman & Hall/CRC Press; 2021.

19. Mackie AS, Islam S, Magill-Evans J, Rankin KN, Robert C, Schuh M, Nicholas D, Vonder Muhll I, McCrindle BW, Yasui Y, et al. Healthcare transition for youth with heart disease: a clinical trial. Heart. 2014;100(14):1113–1118. DOI: 10.1136/heartjnl-2014-305748.

20. Goossens E, Fieuws S, Van Deyk K, Luyckx K, Gewilling M, Budts W, Moons P. Effectiveness of structured education on knowledge and health behaviors in patients with congenital heart disease. J Pediatr. 2015;166(6):1370–1376. DOI: 10.1016/j.jpeds.2015.02.041.

21. Lee S, Lee J, Choi JY. The effect of a resilience improvement program for adolescents with complex congenital heart disease. Eur J Cardiovasc Nurs. 2017;16(4):290–298. DOI: 10.1177/1474515116659836.

22. Hergenroeder AC, Moodie DS, Penny DJ, Wiemann CM, Sanchez-Fournier B, Moore LK, Head J. Functional classification of heart failure before and after implementing a healthcare transition program for youth and young adults transferring from a pediatric to an adult congenital heart disease clinics. Congenit Heart Dis. 2018;13(4):548–553. DOI: 10.1111/chd.12604.

23. Mackie AS, Rempel GR, Kovacs AH, Kaufman M, Rankin KN, Jelen A, Yaskina M, Sananes R, Oechslin E, Dragieva D, et al. Transition intervention for adolescents with congenital heart disease. J Am Coll Cardiol. 2018;71(16):1768–1777. DOI: 10.1016/j.jacc.2018.02.043.

24. Lee MJ, Jung D. Development and effects of a self-management efficacy promotion program for adult patients with congenital heart disease. Eur J Cardiovasc Nurs. 2019;18(2):140–148. DOI: 10.1177/1474515118800099.

25. Gaydos SS, Chowdhury SM, Judd RN, McHugh KE. A transition clinic intervention to improve follow-up rates in adolescents and young adults with congenital heart disease. Cadiol Young. 2020;30(5):633–640. DOI: 10.1017/S1047951120000682.

26. Bushee C, Ginde S, Earing MG, Buelow M, Reinhardt E, Cohen S. Changes in care patterns associated with a transition program in adolescents with congenital heart disease: a single center study. Progress in Pediatric Cardiology. 2021;62:101343. DOI: 10.1016/j.ppedcard.2021.101343.

27. Hwang JH. Effects of an online health management program for adolescents with complex congenital heart disease during their transition to adulthood. Unpublished dissertation. Seoul: Seoul National University; 2022.

28. Mackie AS, Rankin KN, Yaskina M, Gingrich J, Williams E, Schuh M, Kovacs AH, McCrindle BW, Nicholas D, Rempel GR. Transition preparation for young adolescents with congenital heart disease: a clinical trial. J Pediatr. 2022;241:36–41.e2. DOI: 10.1016/j.jpeds.2021.09.053.

29. Bandura A. Self-efficacy: toward a unifying theory of behavioral change. Psychol Rev. 1977;84(2):191–215.

30. McHugh ML. Interrater reliability: the kappa statistic. Biochem Med. 2012;22(3):276–282. DOI: 10.11613/BM.2012.031.

31. Sable C, Foster E, Uzark K, Bjornsen K, Canobbio MM, Connolly HM, Graham TP, Gurvitz MZ, Kovacs A, Meadows AK. Best practices in managing transition to adulthood for adolescents with congenital heart disease: the transition process and medical and psychosocial issue: a scientific statement from the American Heart Association. Circulation. 2011;123(13):1454–1485. DOI: 10.1161/CIR.0b013e3182107c56.

32. Field MJ, Jette AM; Institute of Medicine (US) Committee on Disability in America (editors). The Future of Disability in America. Washington, D.C.: National Academies Press; 2007.

33. Mackie AS, Fournier A, Swan L, Marelli AJ, Kovacs AH. Transition and transfer from pediatric to adult congenital heart disease care in Canada: call for strategic implementation. Can J Cardiol. 2019;35(12):1640–1651. DOI: 10.1016/j.cjca.2019.08.014.

34. Ladouceur M, Iserin L, Cohen S, Legendre A, Boudjemline Y, Bonnet D. Key issues of daily life in adults with congenital heart disease. Arch Cardiovasc Dis. 2013;106(6-7):404–412. DOI: 10.1016/j.acvd.2013.02.004.

35. Kovacs AH, Brouillette J, Ibeziako P, Jackson JL, Kasparian NA, Kim YY, Livecchi T, Sillman C, Kochilas LK; American Heart Association Council on Lifelong Congenital Heart Disease and Heart Health in the Young, et al. Psychological outcomes and interventions for individuals with congenital heart disease: a scientific statement from the American Heart Association. Circulation. 2022;15(8):e000110. DOI: 10.1161/HCQ.0000000000000110.

36. Cocomello L, Dimagli A, Biglino G, Cornish R, Caputo M, Lawlor DA. Educational attainment in patients with congenital heart disease: a comprehensive systematic review and meta-analysis. BMC Cardiovasc Disord. 2021;21(1):549. DOI: 10.1186/s12872-021-02349-z.

37. Westhoff-bleck M, Briest J, Fraccarollo D, Hilfiker-kleiner D, Winter L, Maske U, Busch MA, Bleich S, Bauersachs J, Kahl KG. Mental disorders in adults with congenital heart disease: Unmet needs and impact on quality of life. J Affect Disord. 2016;204:180–186. DOI: 10.1016/j.jad.2016.06.047.

38. Luychx K, Goossens E, Rassart J, Apers K, Vanhalst J, Moons P. Parental support, internalizing symptoms, perceived health status, and quality of life in adolescents with congenital heart disease: influences and reciprocal effects. J Behav Med. 2014;37(1):145–155. DOI: 10.1007/s10865-012-9474-5.

39. Heery E, Sheehan AM, While AE, Coyne I. Experiences and outcomes of transition from pediatric to adult health care services for young people with congenital heart disease: a systematic review. Congenit Heart Dis. 2015;10(5):413–427. DOI: 10.1111/chd.12251.

40. Flocco SF, Lillo A, Dellafiore F, Goossens E. Congenital heart disease: the nursing care handbook. Switzerland: Springer International Publishing; 2019.

41. Eun YM. [Expert column] Adult congenital heart disease patients have nowhere to go. The Dong-A Ilbo. 2023 February 2. Available from https://www.donga.com/news/article/all/20230201/117698338/1. Accessed May 15, 2023.

42. Moons P, De Volder E, Budts W, De Geest S, Elen J, Waeytens K, Gewillig M. What do adult patients with congenital heart disease know about their disease, treatment, and prevention of complications? A call for structured patient education. Heart. 2001;86(1):74–80. DOI: 10.1136/heart.86.1.74.

43. Sawicki GS, Lukens-Bull K, Yin X, Demars N, Huang IC, Livingood W, Reiss J, Wood D. Measuring the transition readiness of youth with special healthcare needs: validation of the TRAQ—Transition Readiness Assessment Questionnaire. J Pediatr Psychol. 2011;36(2):160–171. DOI: 10.1093/jpepsy/jsp128.

44. De Lima Campos EF, Perin L, Assmann M, Lucchese F, Pellanda LC. Knowledge about the disease and the practice of physical activity in children and adolescents with congenital heart disease. Arq Bras Cardiol. 2020;114(5):786–792. DOI: 10.36660/abc.20180417.

45. Kwon SJ, Im YM. Sexual health knowledge and needs among young adults with congenital heart disease. PLoS One. 2021;16(5):e0251155. DOI: 10.1371/journal.pone.0251155.

46. Van Deyk K, Pelgrims E, Troost E, Goossens E, Budts W, Gewillin M, Moons P. Adolescents’ understanding of their congenital heart disease on transfer to adult-focused care. Am J Cardiol. 2010;106(12):1803–1807. DOI: 10.1016/j.amjcard.2010.08.020.

47. Ebbinghaus H. Memory: a contribution to experimental psychology (reprint of 1913 Edition). Manchester, CT: Martino Fine Books; 2011.

48. Riegel B, Lee CS, Dickson VV, Carlson B. An update on the self-care of heart failure index. J Cardiovasc Nurs. 2009;24(6):485–497. DOI: 10.1097/JCN.0b013e3181b4baa0.

49. Van Achterberg T, Huisman-de Waal GG, Ketelaar NA, Oostendorp RA, Jacobs JE, Wollershein HC. How to promote healthy behaviours in patients? An overview of evidence for behaviour change techniques. Health Promot Int. 2011;26(2):148–162. DOI: 10.1093/heapro/daq050.

50. García-Rodríguez F, Raygoza-Cortez K, Moreno-Hernandez L, García-Pérez R, Lopez LE, Arana-Guajardo AC, Jáquez-Quintana JO, Villarreal-Treviño AV, De la O-Cavazos ME, Rubio-Pérez N. Outcomes of transitional care programs on adolescent chronic inflammatory systemic diseases: systematic review and meta-analyses. Pediatr Rheumatol Online J. 2022;20(1):15. DOI: 10.1186/s12969-022-00670-1.

51. Bassareo PP, Chessa M, Di Salvo G, Walsh KP, Mcmahon CJ. Strategies to aid successful transition of adolescents with congenital heart disease: a systematic review. Children. 2023;10(3):423. DOI: 10.3390/children10030423.

52. Bertoletti, J., Marx, G. C., Hattge, S. P., & Pellanda, L. (2015). Health-related quality of life in adolescents with congenital heart disease. Cardiology in the Young, 25(3), 526–532. DOI: 10.1017/S1047951114000304.

53. Khajali Z, Sayyadi A, Ansari Z, Aliramezany M. Quality of life in adult patients with congenital heart disease: results of a double-center study. Front Psychiatry. 2023;13:1062386. DOI: 10.3389/fpsyt.2022.1062386.

54. Liu HC, Chaou CH, Lo CW, Chung HT, Hwang MS. Factors affecting psychological and health-related quality-of-life status in children and adolescents with congenital heart diseases. Children. 2022;9(4):578. DOI: 10.3390/children9040578.

55. Xu C, Su X, Ma S, Shu Y, Zhang Y, Hu Y, Mo X. Effects of exercise training in postoperative patients with congenital heart disease: a systematic review and meta-analysis of randomized controlled trials. J Am Heart Assoc. 2020;9(5):e013516. DOI: 10.1161/JAHA.119.013516.

